# Antimicrobial resistance patterns among urinary isolates from long-term care residents in Canada: findings from an AMRNet pilot

**DOI:** 10.64898/2026.10.29.26366712

**Authors:** Chanchal Yadav, Kanchana Amaratunga, Amrita Bharat, Kevin Afra, Kevin A. Brown, Connor Chato, Nick Daneman, Bradley Langford, Gerry McDonald, Xiaofeng Ding, Chris Norgaard, Meredith Faires, Linda Hoang, Pamela Kibsey, Eunsun Lee, Annie Mak, Jessica Minion, Peter Daley, Wallis Rudnick, AMRNet

## Abstract

**Background:** Antimicrobial resistance is a growing concern in long-term care facilities (LTCs), where residents are often older, have chronic health conditions, and are frequently prescribed antimicrobials. Historically, antimicrobial resistance data from Canadian LTCs have been limited. This study explores the use of lab-based surveillance data from the Public Health Agency of Canada’s AMRNet program to describe antimicrobial resistance in LTCs.

**Methods:** Antimicrobial susceptibility data for clinical bacterial specimens were submitted from public and private laboratories across Canada. Where feasible, LTC patients were identified by submitting jurisdictions.

Data on urinary isolates from identified LTC patients were analysed using the first isolate per patient per species per calendar year. The data were described by distribution of age group, sex, species and antimicrobial susceptibility. Antimicrobials were classified by the World Health Organization’ s ‘Access, Watch and Reserve’ (AWaRe) framework and a weighted-incidence syndromic combination antibiogram was constructed.

As part of AMRNet program development, in some jurisdictions, only certain specimen types or organisms were submitted. Not all jurisdictions participated every year. Due to privacy precautions, some values rely on estimations.

**Results:** From 2016–2024, AMRNet received data on 216,524 isolates from five provinces (BC, NL, ON, PEI, SK). Most isolates (98%) were from patients aged ≥60, with those ≥80 years representing 80%; females predominated across all age groups.

Urine represented >99% of submitted data. The most common organisms identified from urinary isolates were *Escherichia coli* (51%), *Klebsiella pneumoniae* (14%) and *Proteus mirabilis* (12%). Non-susceptibility varied across organisms: *E. coli* demonstrated higher non-susceptibility to several commonly used agents including fluoroquinolones (44%) and third-generation cephalosporins (18%) while *K. pneumoniae* displayed high non-susceptibility to nitrofurantoin (56%). Among *P. mirabilis,* 26% were non-susceptible to ciprofloxacin. Aminoglycoside and carbapenem resistance were low. When antimicrobials were grouped by WHO AWaRe categories, more than half of *E. coli* isolates were non-susceptible to at least one Watch antimicrobial.

**Interpretation:** AMRNet lab-based surveillance data can help describe national AMR patterns among LTC residents. Although limitations exist and identifying LTC patients in lab-based datasets can be challenging in some jurisdictions, these data provide a feasible and cost-effective means of tracking national resistance trends among an at-risk population.

## Introduction

Antimicrobial resistance (AMR) has emerged as a major threat to global health, reducing the effectiveness of antimicrobials used to treat both routine and severe infections (1–4). As microorganisms develop mechanisms that enable them to withstand antimicrobial therapy, infections become harder to treat, resulting in longer illness duration, increased healthcare utilization and costs, and higher mortality worldwide (5–7).

Residents of long-term care (LTC) facilities are particularly vulnerable to the consequences of AMR due to a combination of factors, including advanced age, multimorbidity, high rates of antimicrobial use, and frequent interactions with healthcare services and on-site medical care, creating repeated opportunities for exposure to resistant organisms (4, 8–10). Residents in long term care (LTC) facilities experience a range of infections; however, urinary tract infections (UTIs) are consistently among the most commonly reported, accounting for approximately 20% to over 35% of all infections in some facilities (11–14). UTIs are a major cause of antimicrobial use in this population, comprising more than one third of all antibiotic prescriptions (15–18). Urine cultures are frequently obtained in the absence of clinical symptoms, leading to increased likelihood of misclassifying asymptomatic bacteriuria as UTIs and contributing to unnecessary antimicrobial use (19).

In Canada, national AMR surveillance has historically focused on acute-care facilities, with comparatively limited antimicrobial susceptibility data available from LTC facilities (20), representing an important gap in understanding resistance trends. Strengthening surveillance is essential to inform stewardship and infection-prevention efforts in these facilities. To address this gap, the Public Health Agency of Canada’s (PHAC’s) AMRNet surveillance program initiated a pilot project to analyse susceptibility data originating from LTC facilities. Established in 2021, AMRNet is a national laboratory-based AMR surveillance initiative led by the PHAC’s National Microbiology Laboratory in collaboration with provincial and territorial public health organizations and laboratories across the country (21). As of April 2026, AMRNet collects both isolate-level and aggregate clinical antimicrobial susceptibility testing (AST) data from both public and private laboratories in eight participating provinces and territories. Building on this national infrastructure, the pilot project described here provides findings on antimicrobial non-susceptibility patterns in urinary bacterial isolates collected from LTC facilities across participating provinces and territories. As AMRNet data includes contributions from approximately 1,000 LTC facilities, representing about half of the approximately 2,000 LTC facilities in Canada, these results offer an unprecedented view of AMR in this sector (22).

## Materials and Methods

### Data sources and study population

This pilot project analysed clinical AST data from LTC facilities submitted to the Canadian AMRNet surveillance program by participating provinces and territories from 2016–2024. AMRNet, described elsewhere (21), includes antimicrobial susceptibility testing data from bacterial and fungal pathogens from public and private clinical laboratories across Canada. Along with susceptibility test results, AMRNet collects information on specimen type, as well as patients’ sex, age group, setting and geographic location. Screening and surveillance specimens are excluded.

LTC-associated isolates were identified by submitted jurisdictions where feasible. Identifiable LTC data were available from five provinces and included both isolate-level (4 jurisdictions) and aggregate submissions (1 jurisdiction).

As a developing surveillance system, not all jurisdictions participated in all years of surveillance. Participation in LTC surveillance increased from 3 jurisdictions for the 2017/18 datasets to 5 jurisdictions for the 2023/24 datasets. While AMRNet aims to collect all susceptibility test results from all geographic regions in a province, in 2/5 provinces, some regions were not yet onboarded during the study period, resulting in estimated coverage of approximately 60% and 85%, respectively.

#### Isolate-level data

Raw data submitted by participating provinces were standardized centrally by AMRNet using a custom R-based pipeline and the AMR R package (23) prior to analyses.

#### Aggregate data

One jurisdiction submitted aggregate rather than individual-level data. Aggregate data consisted of counts of susceptible (S) and non-susceptible (NS) results for each organism-antimicrobial combination, stratified by available covariates (specimen source, sex, age group, setting, region, and year). These data included only the first isolate per organism per patient in a calendar year and were limited to AST results reported to clinicians. For privacy, some small cell counts were suppressed and imputed with expected mean values.

### Laboratory methods

Organism identification and AST were performed by laboratories according to routine diagnostic procedures in each jurisdiction. All jurisdictions report the routine use of CLSI methods and breakpoints in their jurisdictions although a small proportion of laboratories use alternative methods (e.g., EUCAST).

### Data analysis procedures

Descriptive analyses were conducted to summarize organism distribution, demographic patterns, and antimicrobial susceptibility patterns. Duplicate isolates were removed according to Clinical and Laboratory Standards Institute (CLSI) guidance (24) with only the first clinical isolate per patient per organism per calendar year included.

Antimicrobial non-susceptibility analyses, including AWaRe, and weighted-incidence syndromic combination antibiogram (WISCA) estimates were further restricted using AMRNet reporting criteria. To ensure comparability and reduce potential bias arising from selective testing and variation in laboratory reporting practices across jurisdictions, data from a jurisdiction were included for a given organism-antimicrobial combination only when ≥85% of isolates for that organism had a susceptibility result available for the antimicrobial (tested or reported). Following application of this jurisdiction-level criterion, organism-antimicrobial combinations were reported only when:

1. Data for the combination were contributed by at least three participating provinces or territories; and
2. At least 30 isolates were available for analysis (as per CLSI recommendations).

Data submitted that did not meet the AMRNet reporting criteria above were excluded from susceptibility estimates.

Non-susceptibility was defined as intermediate or resistant susceptibility test results and was calculated as:

(Number of non-susceptible results ÷ Total susceptibility test results reported) x 100

Because AST and reporting practices differed across jurisdictions, the number of isolates available for analysis varied by organism-antimicrobial combination.

Consequently, denominators differ between organisms and antimicrobials throughout the susceptibility analyses. Susceptibility estimates were based on data as submitted by participating jurisdictions; missing susceptibility results were not imputed, and no additional interpretative or expert rules were applied centrally.

To contextualize susceptibility patterns across antimicrobials, results were categorized according to the World Health Organization Access, Watch, and Reserve (AWaRe) classification. Only isolate-level data were included in this portion of the analyses.

A weighted-incidence syndromic combination antibiogram (WISCA) was constructed using the three most common urinary pathogens in the AMRNet dataset, representing over 77**%** of all urine isolates. The WISCA estimates probability that empiric antimicrobial therapy will have in vitro activity against a urinary pathogen and complements the organism specific susceptibility analyses by reflecting the clinical syndrome and pathogen frequency rather than individual organisms (25) Antimicrobial-organism combinations classified as intrinsically resistant according to CLSI M100 definitions were retained and considered resistant in the overall susceptibility estimates.

### Ethics approval

As data were collected and analyzed for routine public health surveillance purposes at the Public Health Agency of Canada, research ethics board review was not required.

## Results

### Participating provinces and territories and isolate counts

Five jurisdictions (BC, NL, ON, PEI, SK) submitted data in which LTC facilities could be identified (Figure 1). In total, 216,524 urine isolates were reported from LTC facilities.

**Figure 1:**
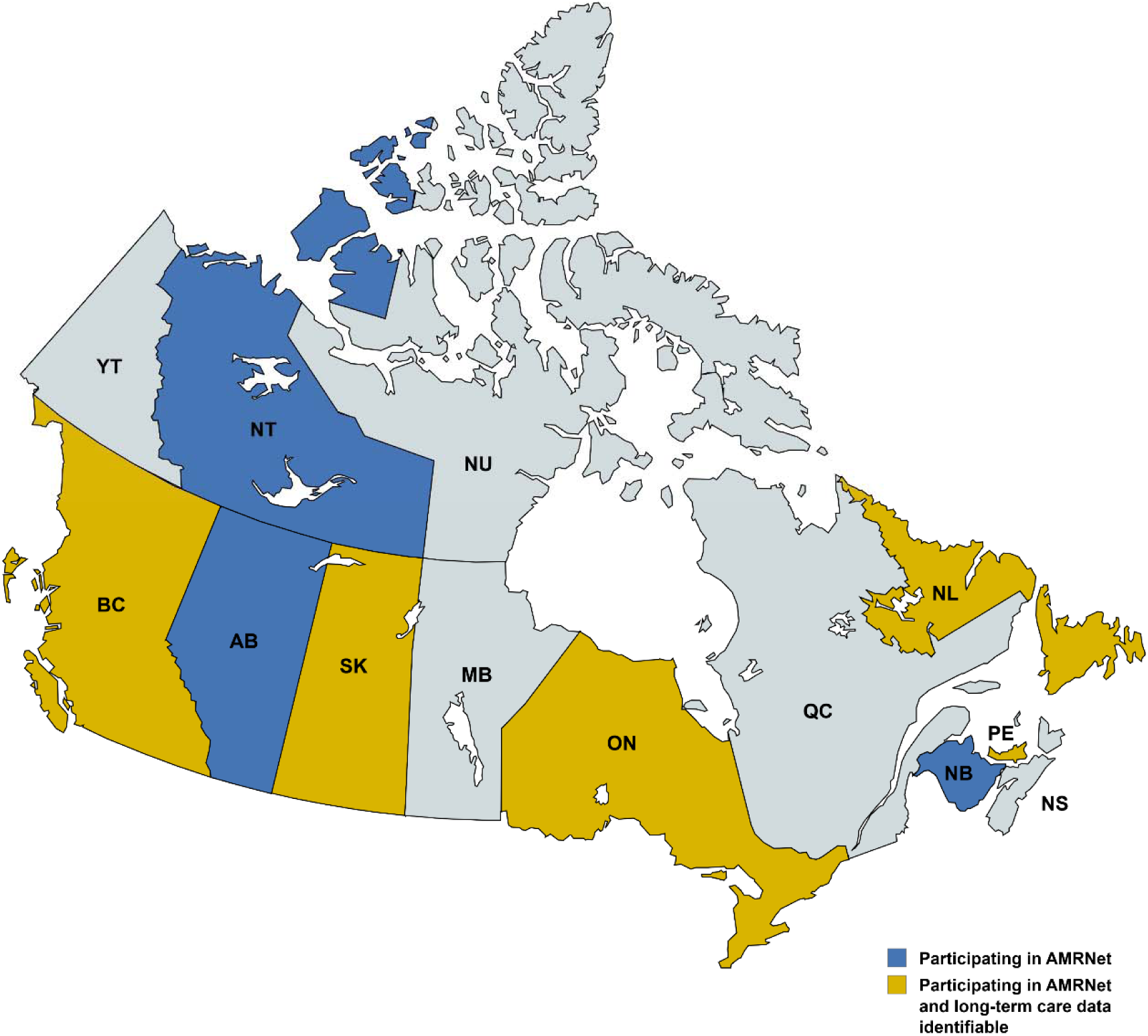
AMRNet participation and submission status for long-term care settings, 2024. Provinces/Territories: BC=British Columbia, AB=Alberta, SK=Saskatchewan, MB=Manitoba, ON=Ontario, QC=Quebec, NB=New Brunswick, NS=Nova Scotia, PE=Prince Edward Island, NL=Newfoundland & Labrador; YT=Yukon, NT=Northwest Territories, NU=Nunavut.

Following CLSI-based deduplication and exclusion of isolates lacking susceptibility interpretations, 214,360 urinary isolates were included in the analysis.

### Age and sex distribution of urinary isolates

Age and sex information was available for 97% of urinary isolates. The distribution of urinary isolates by age group and sex is presented in Supplementary Figure 1.

Individuals aged ≥ 80 years accounted for the largest proportion of urinary isolates (80%) in long-term care facilities. Across all age groups, females consistently contributed the majority of isolates (75% of isolates overall).

### Organism distribution

The distribution of organisms among urinary isolates is shown in Figure 2. *Escherichia coli* accounted for 51% of total isolates, followed by *Klebsiella pneumoniae* (14%) and *Proteus mirabilis* (12%). Together, these three Enterobacterales comprised >75% of urinary isolates. *Enterococcus faecalis,* other *Enterococcus* species and *Staphylococcus aureus* were among the most common gram-positive organisms and represented approximately 2–3% of urinary isolates each. Among *S. aureus* isolates, 32% were methicillin-resistant (MRSA) during 2022–2024.

**Figure 2:**
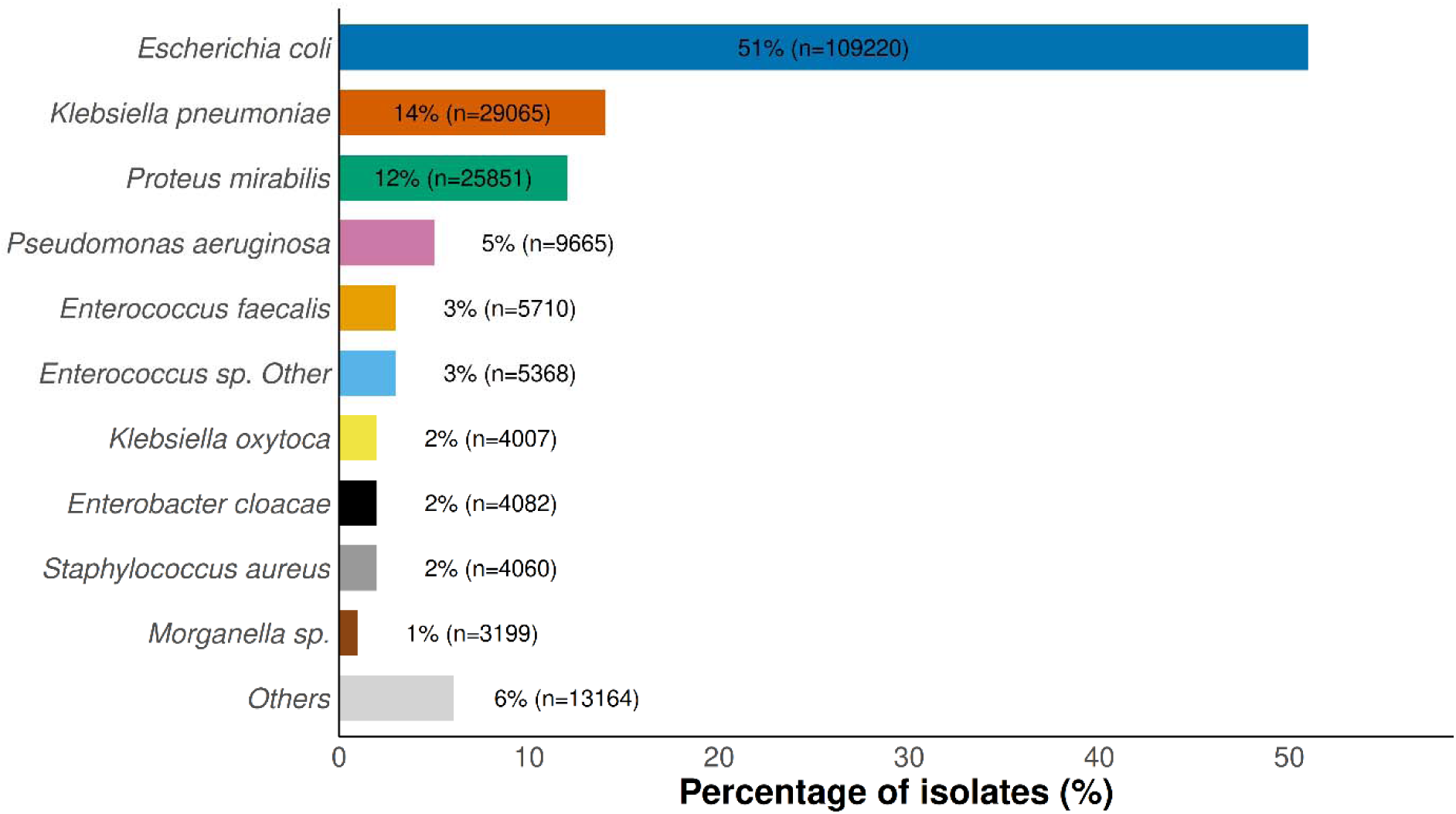
Ten most frequently reported organisms from urinary specimens from long-term care facilities, and all other organisms combined., AMRNet 2016–2024.

### Antimicrobial non-susceptibility among urinary isolates

The percentage of non-susceptible isolates for select antimicrobial-organism combinations is presented in Figure 3. As Enterobacterales comprised the majority of urinary isolates, non-susceptibility patterns were examined for the top three most frequently identified Enterobacterales species: *E.coli, K. pneumoniae and P. mirabilis*.

**Figure 3:**
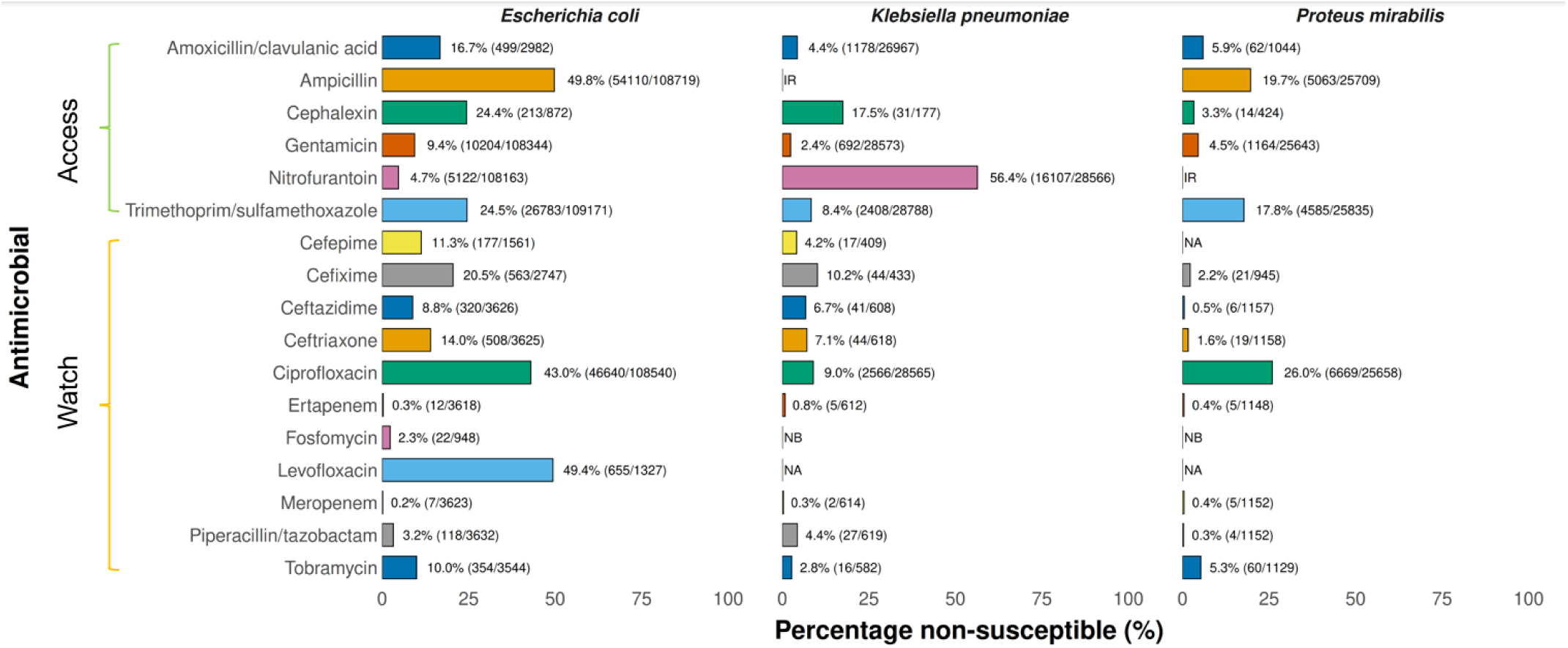
Percentage non-susceptible by antimicrobial and organism from urinary specimens from long-term care facilities, AMRNet 2016–2024. NA = Not Available (did not meet reporting criteria); IR= Intrinsically Resistant as per CLSI M100; NB = No Breakpoint defined.

Among the three most common Enterobacterales, notable differences in non-susceptibility patterns were observed. *E. coli* demonstrated much lower non-susceptibility to nitrofurantoin (4.7%) than *K. pneumoniae* (>50%). In contrast, non-susceptibility to trimethoprim-sulfamethoxazole, amoxicillin-clavulanate, and fluoroquinolones was higher in *E. coli* than in *K. pneumoniae* (24.5%, 16.7%, and 43.0% vs. 8.4%, 4.4%, and 9.0%, respectively). *Proteus mirabilis* demonstrated intermediate non-susceptibility to ciprofloxacin (26.0%), higher than *K. pneumoniae* but lower than *E. coli*. Carbapenem non-susceptibility remained rare in all the three Enterobacterales (<1%), while non-susceptibility to aminoglycosides (gentamicin and tobramycin) was low across all organisms (≤10%).

Among the three most common Enterobacterales, non-susceptibility to at least one third-generation cephalosporin (ceftriaxone, cefotaxime, or ceftazidime) was highest in *E. coli* (18.3%), followed by *K. pneumoniae* (12.0%). In contrast, non-susceptibility was rare in *P. mirabilis* (1.3%).

When grouped by WHO AWaRe categories, non-susceptibility differed among the three predominant Enterobacterales (Figure 4, Supplementary Table 1). *E. coli* showed the highest non-susceptibility to both Access (56.6%) and Watch (49.4%) antibiotics compared with *K. pneumoniae* (48.0% and 13.8%, respectively). *P. mirabilis* demonstrated intermediate levels of non-susceptibility to Access (35.8%) and Watch (36.6%) antibiotics. No Reserve antimicrobials met AMRNet’s reporting criteria.

**Figure 4:**
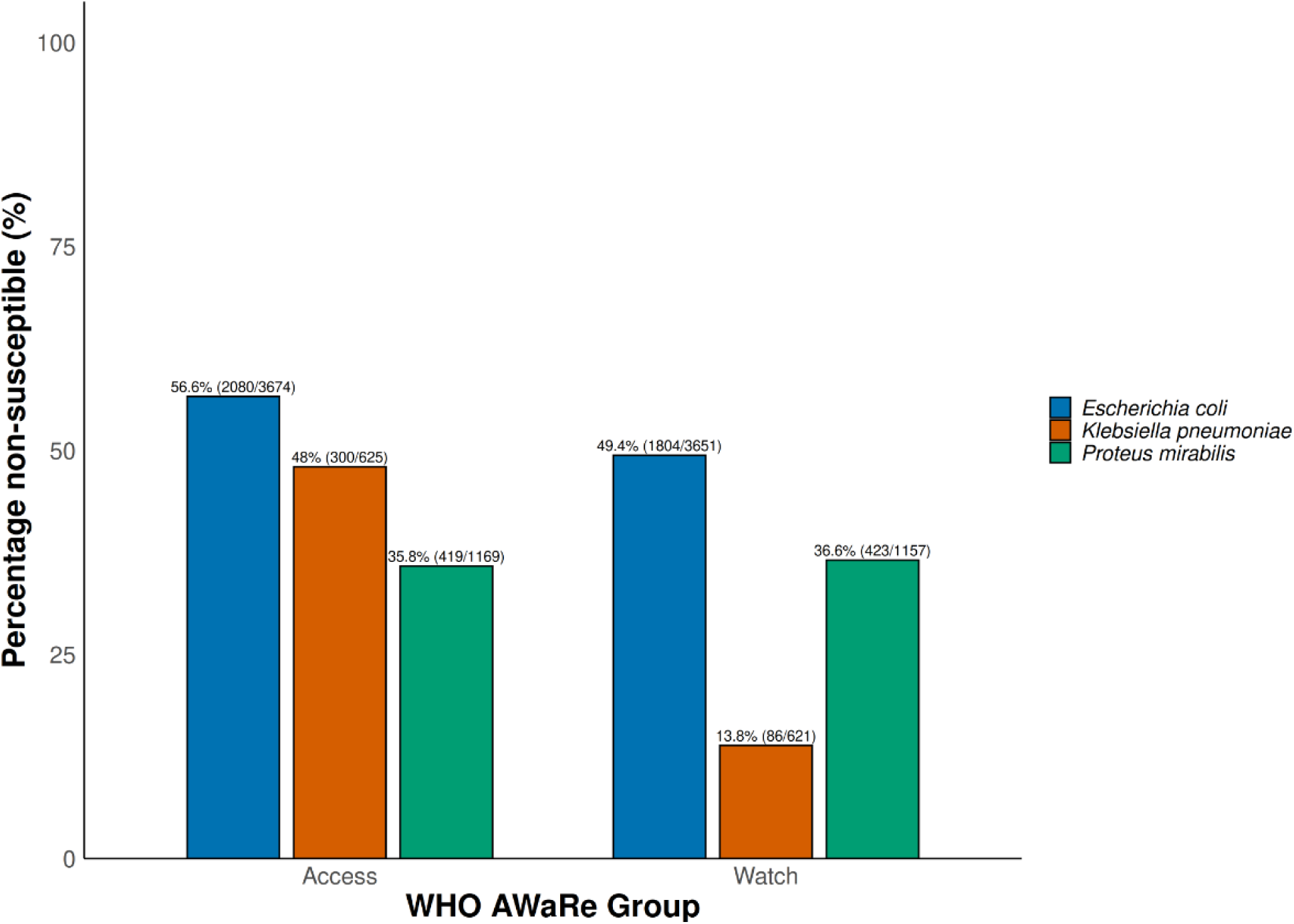
Percentage non-susceptible to ≥1 World Health Organization Access or Watch category antibiotics among urine isolates from long-term care settings, AMRNet 2022– 2024. Four provinces represented; aggregate data excluded.

#### Weighted-incidence syndromic combination antibiogram

A WISCA was developed using the three most frequent organisms from urinary specimens (*Escherichia coli*, *Klebsiella pneumoniae*, and *Proteus mirabilis*) (25) (Table 1). Susceptibility among urine isolates was found to be highest for ceftriaxone (91.2%) and amoxicillin/clavulanic acid (89.0%). Susceptibility was approximately 80% for cephalexin (81.4%), trimethoprim/sulfamethoxazole (81.2%), and nitrofurantoin (80.6%). Lower susceptibility was observed for ciprofloxacin (65.9%) and ampicillin (47.5%).

**Table 1:**
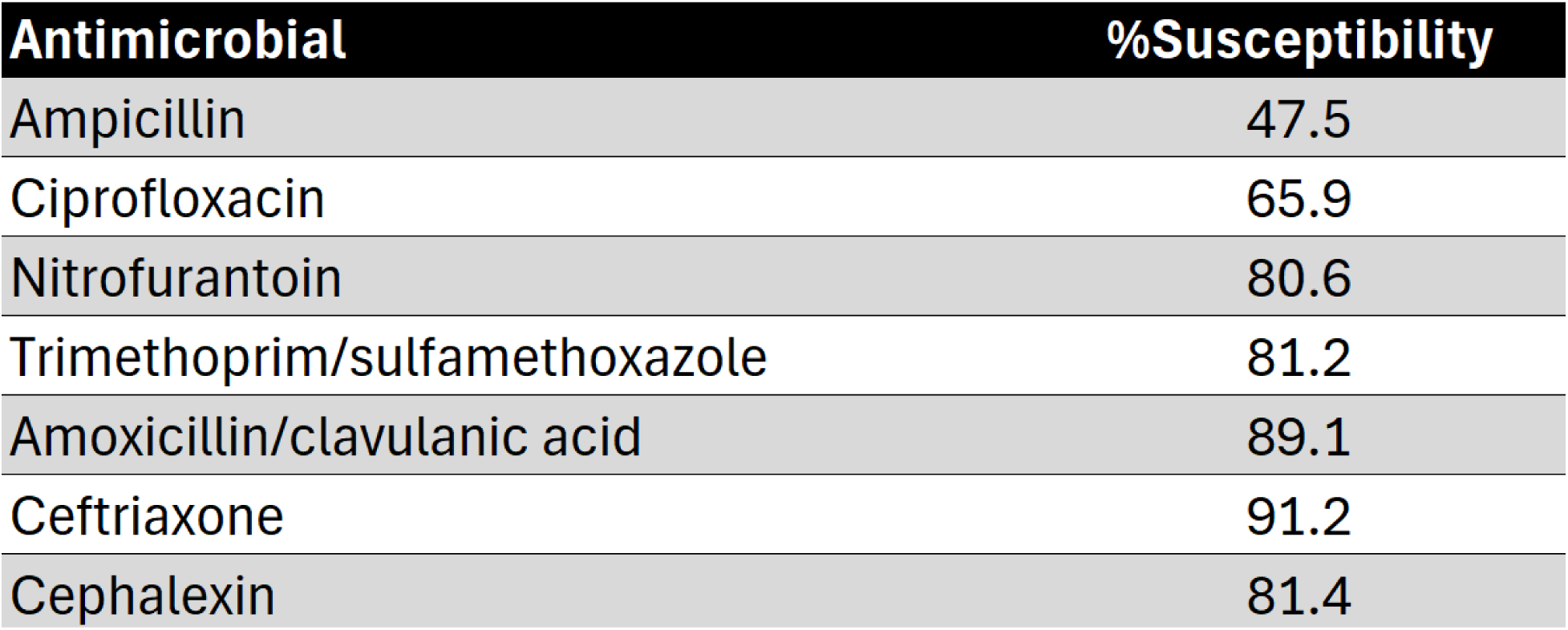
Weighted-incidence syndromic combination antibiogram illustrating the percentage of susceptible urinary isolates for the three most common gram negative organisms (*E. coli, K. pneumoniae, and P. mirabilis*) reported from long-term care facilities, AMRNet 2016–2024. Included organisms represented 77% of all reported urinary specimens and approximately 83% of gram-negatives.

| **Antimicrobial** | **%Susceptibility** |
|---|---|
| Ampicillin | 47.5 |
| Ciprofloxacin | 65.9 |
| Nitrofurantoin | 80.6 |
| Trimethoprim/sulfamethoxazole | 81.2 |
| Amoxicillin/clavulanic acid | 89.1 |
| Ceftriaxone | 91.2 |
| Cephalexin | 81.4 |

## Discussion

To our knowledge, this pilot project presents the first national-level description of antimicrobial non-susceptibility patterns among urinary bacterial isolates from Canadian LTC facilities, establishing initial benchmarks in this sector. Using routinely collected laboratory data submitted through AMRNet, this analysis demonstrates the feasibility and value of coordinated, multijurisdictional lab-based surveillance to inform stewardship and monitor resistance trends in LTC facilities.

This predominance likely reflects the high burden of urinary tract infections, which are among the most common infections and indications for antimicrobial use in LTC populations. It may also reflect substantial variation in urine culturing and diagnostic practices that persists, in some studies, even after accounting for resident characteristics and may represent opportunities for antimicrobial stewardship (9, 13, 16-18(26, 27). Together, these factors likely contributed to the predominance of urinary isolates in our dataset. These findings underscore the importance of antimicrobial stewardship efforts aimed at optimizing urine culture ordering and reducing unnecessary treatment of asymptomatic bacteriuria.

Consistent with prior reports, the Gram-negative organisms *E.coli, K. pneumoniae* and *P. mirabilis* were the most frequently identified organisms and represented more than three-quarters of all isolates, highlighting the predominance of Enterobacterales in this population (Figure 3) (11, 28, 29). Among gram-positive organisms, *Enterococcus faecalis,* other *Enterococcus* species and *S. aureus* represented a smaller proportions (2-3% each) of urine isolates; nearly one-third of *S. aureus* isolates were found to be methicillin-resistant among LTC isolates compared to one-fifth in overall AMRNet surveillance including all facilities (30). MRSA bacteriuria frequently results from colonization or contamination rather than true urinary infection, particularly among older adults and those residing in long-term care facilities (31). Studies have previously reported that *S. aureus* in urine is often nonspecific and may not reflect invasive disease, which is better assessed through bloodstream surveillance (32).

In this study, non-susceptibility patterns for *E. coli*, *K. pneumoniae* and *P. mirabilis* broadly aligned with published reports from LTC facilities in Canada and the United States. Although direct comparison are limited by differences in study design, laboratory practices and populations, these prior studies similarly report high susceptibility to nitrofurantoin and elevated non-susceptibility to trimethoprim-sulfamethoxazole and, notably, to fluoroquinolones in long-term care settings (11, 28, 29, 33). Consistent with known species characteristics, *K. pneumoniae* had relatively high non-susceptibility to nitrofurantoin in this study, reflecting its reduced susceptibility to this antimicrobial.

LTC facilities often have lower proportions of non-susceptible isolates compared with inpatient facilities (28, 34, 35) but higher proportions than community (36, 37). Consistent with this, we found a generally higher proportion of non-susceptibility among the isolates from LTC facilities compared to general AMRNet surveillance (all settings combined). For instance, 43% of E. *coli* urinary isolates from LTC facilities were non-susceptible to ciprofloxacin compared with 30% in general AMRNet surveillance. This is likely due to the high proportion of isolates from outpatient/community settings in combined surveillance with lower rates of non-susceptibility.

Non-susceptibility to fluoroquinolones exceeded 40% in *E. coli.* This is consistent with prior studies from Canada and the United States reporting non susceptibility often exceeding 30%, with some LTC facilities reporting rates above 60% (11, 28, 35). This has direct implications for empiric therapy in these facilities. Fluoroquinolones are not recommended as first-line therapy for UTI treatment, yet they continue to comprise a substantial proportion of antibiotic use in LTC facilities, representing approximately one-fifth to one-third of all prescriptions despite reduced susceptibility and well documented risks that include, for example, development of *Clostridioides difficile* infection (9, 17, 38–44).

Non-susceptibility to third-generation cephalosporins varied among the predominant Enterobacterales and was highest in *E. coli* (18.3%), followed by *K. pneumoniae* (12.0%), while remaining uncommon in *P. mirabilis* (1.8%).. Increasing rates of non-susceptibility to third-generation cephalosporins among Enterobacterales has been observed among urinary isolates in LTC facilities (33, 45) as well as in AMRNet data from all settings (30). Resistance to third-generation cephalosporins is often accompanied by resistance to other agents, such as fluoroquinolones, limiting empiric treatment options for complicated or more serious UTIs (46–48).

In contrast to fluroquinolones and third-generation cephalosporins, non-susceptibility to carbapenems and aminoglycosides remained low across *E. coli, K. pneumoniae*, and *P. mirabilis*. Although not restricted to long-term care facilities, this is similar to what has been reported from urine cultures in Ontario (28), from a surveillance study of Canadian hospitals (49) and from AMRNet data all settings (30). Carbapenems and aminoglycosides are not routinely used in LTC facilities; high susceptibility indicates that these agents remain largely preserved among the predominant urinary Enterobacterales.

The AWaRe classification, introduced by the WHO in 2017, was designed as a stewardship framework to support monitoring of antimicrobial use and emerging resistance (49, 50). When antibiotics were grouped by WHO’s Access, and Watch AWaRe categories, a substantial proportion of urinary isolates demonstrated non-susceptibility to Access and Watch antibiotics (Figure 4). Non-susceptibility to

Access antimicrobials was highest in *E. coli and K. pneumoniae*, while non-susceptibility to Watch antimicrobials was most pronounced in *E. coli* followed by *P. mirabilis* and was lower in *K. pneumoniae.* These findings highlight resistance to both first-line and broader-spectrum antimicrobials used to treat UTIs. No antimicrobials from the “Reserve” category met the reporting criteria for the selected organisms in the AMRNet’s LTC dataset.

A weighted incidence syndromic combination antibiogram (WISCA) was also developed using the three most frequently isolated urinary pathogens, integrating organism prevalence with antimicrobial susceptibility results (Table 1). IDSA aligned guidance recommends nitrofurantoin, trimethoprim sulfamethoxazole, and fosfomycin as preferred empiric agents for uncomplicated urinary tract infections, with broader agents such as third or fourth generation cephalosporins or fluoroquinolones reserved for more complicated presentation (50, 51). The WISCA results broadly reflect these principles: nitrofurantoin (80%) and trimethoprim/sulfamethoxazole (81%) demonstrate moderately high predicted susceptibility. When broader coverage is required, high susceptibility to third generation cephalosporins (91%) supports their reliability.

These findings provide the first multi-jurisdictional overview of antimicrobial resistance in Canadian long-term care facilities, establishing a baseline for future surveillance. The AMRNet dataset represents approximately 1,000 LTC facilities across Canada, based on provincial and territorial LTC counts from the Canadian Institute for Health Information (CIHI), providing a strong foundation for future surveillance. As participation expands and LTC-specific data capture improves, AMRNet will enable more robust tracking of temporal trends, early identification of emerging resistance, and support for evidence-based decision-making and interventions.

Several limitations should be acknowledged. First, participation varied across jurisdictions and years, with not all jurisdictions contributing data throughout the entire study period. Second, while AMRNet aims to collect data on all organisms, specimen types and antimicrobial tested, as part of program development, not all jurisdictions were able to submit all combinations in every year. In addition, AMRNet can only collect data on what is tested and/or what is reported and antimicrobial susceptibility testing practices varied across jurisdictions and over time. To improve comparability and minimize potential bias, AMRNet applied predefined reporting thresholds, including minimum isolate counts and coverage criteria. Data were analysed as reported by jurisdictions. No centralized interpretation of susceptibility test results, including the application of breakpoints or expert rules, was undertaken, and no statistical imputation was performed for missing organism-antimicrobial combinations. Despite these limitations, this pilot establishes a critical foundation for national AMR surveillance in long-term care facilities and, highlights both the feasibility and value of coordinated multi-jurisdictional surveillance efforts.

Strengthening antimicrobial stewardship remains essential to addressing antimicrobial resistance in long-term care facilities. Efforts that support appropriate empiric antimicrobial selection, limit unnecessary urine culture testing, and discourage treatment of asymptomatic bacteriuria are particularly important in these facilities, where residents face high antibiotic exposure and are especially vulnerable to adverse effects and the consequences of resistance (43, 52). Sustained national surveillance efforts will be key for guiding stewardship initiatives and ensuring that Canadian long-term care facilities can respond effectively to the evolving antimicrobial resistance landscape.

## Data Availability

The data that support the findings of this study are available from the Public Health Agency of Canada but restrictions apply to the availability of these data because they were used under agreements with participating jurisdictions and are therefore not publicly available.

## Acknowledgements

We gratefully acknowledge the contribution of the laboratory staff, medical microbiologists, infectious disease physicians, epidemiologists and coordinators at each participating laboratory and public health organization.

We are additionally grateful for the ongoing support of the Computational and Operational Genomics (COG) section at the National Microbiology Laboratory.

---

**Supplementary Figure 1:**
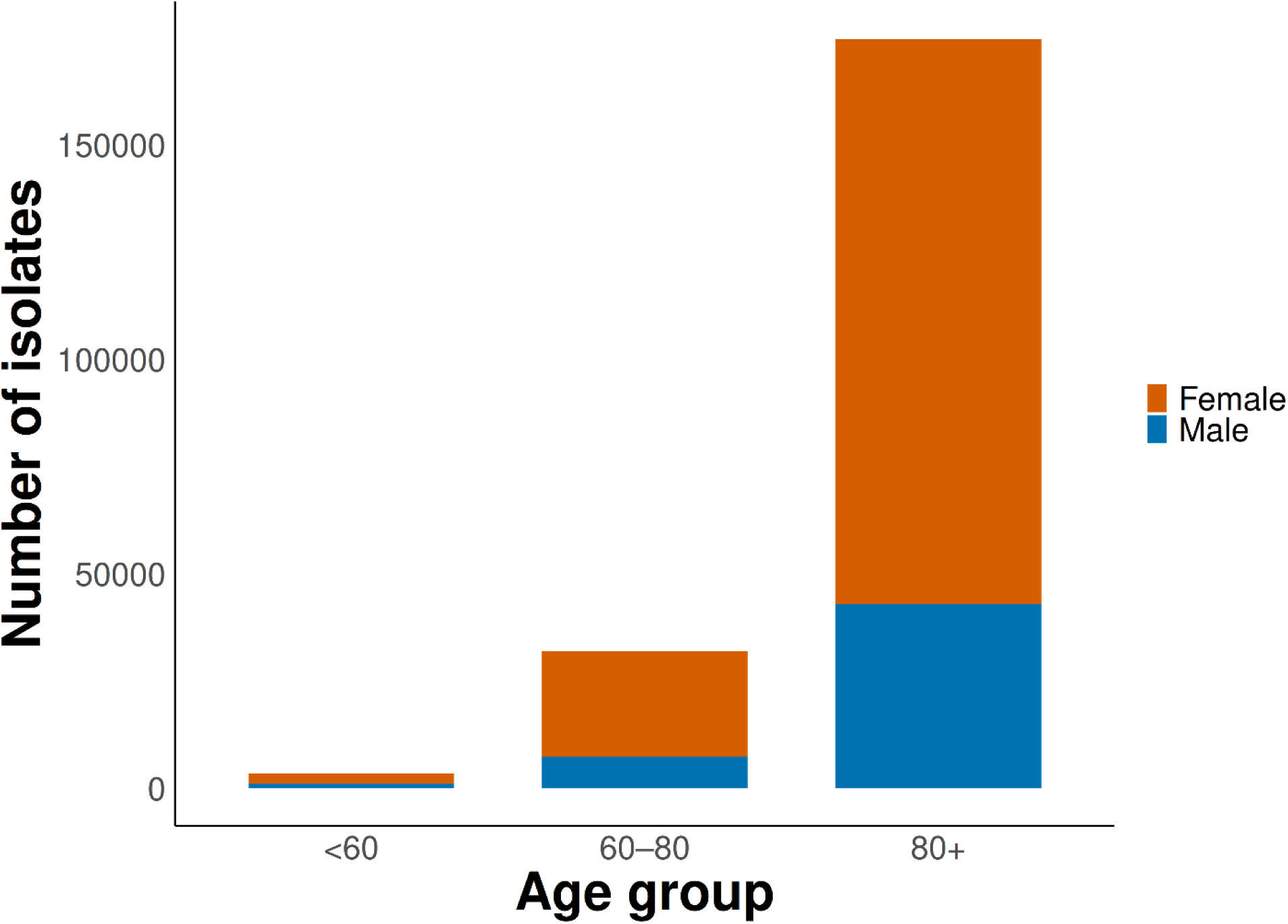
Distribution of urine isolates from long-term care facilities by age-group and sex, AMRNet 2016–2024.

**Supplementary Table 1:**
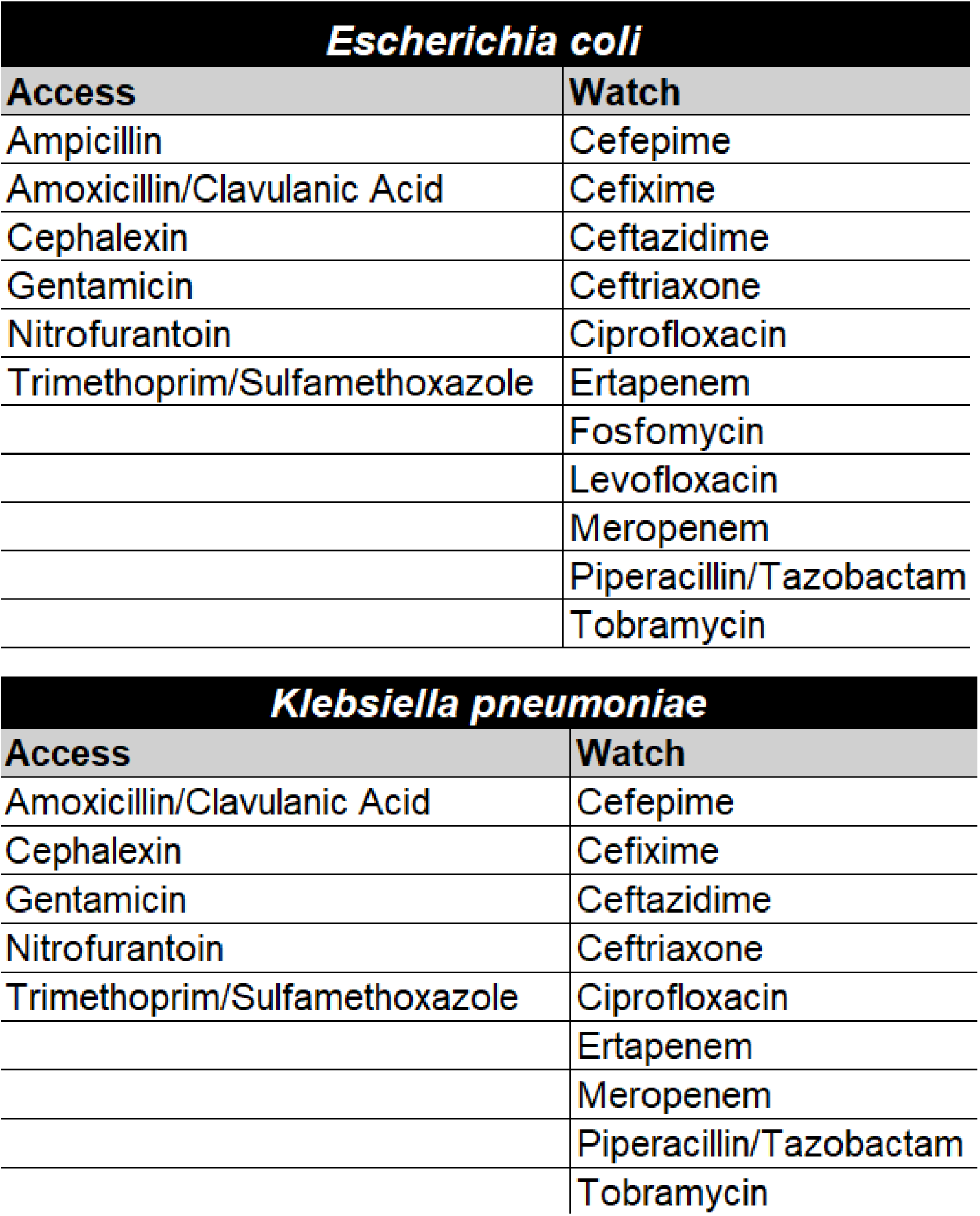
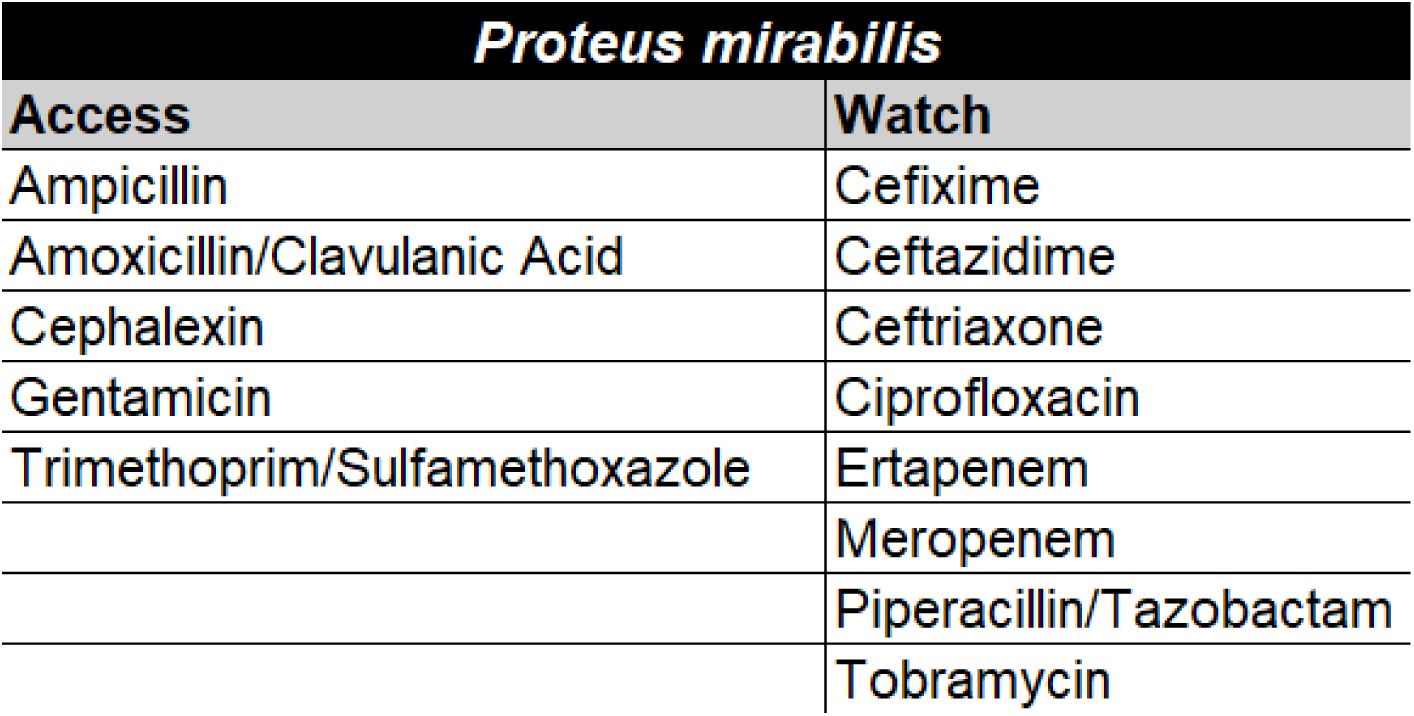
WHO’s Access and Watch category antibiotics underlying the percentage non susceptible estimates for E.coli, K. pneumoniae and P. aeruginosa in Figure 4. Combinations that are intrinsically resistant and those not meeting AMRNet reporting criteria are excluded.

